# Cost-utility analysis of current COVID-19 vaccination program recommendations in Canada

**DOI:** 10.1101/2024.12.13.24318988

**Authors:** Alison E. Simmons, Rafael N. Miranda, Michael W.Z. Li, Gebremedhin B. Gebretekle, Min Xi, Marina I. Salvadori, Bryna Warshawsky, Eva Wong, Raphael Ximenes, Melissa K. Andrew, Sarah Wilson, Matthew Tunis, Ashleigh R. Tuite

**Affiliations:** Centre for Immunization Surveillance and Programs, Public Health Agency of Canada, Ottawa, ON, Canada; Dalla Lana School of Public Health, University of Toronto, Toronto, ON, Canada; Public Health Risk Sciences Division, National Microbiology Laboratory, Public Health Agency of Canada, Guelph, ON, Canada; Department of Mathematics and Statistics, McMaster University, Hamilton, ON, Canada; Department of Pediatrics, McGill University, Montreal, QC, Canada; Department of Epidemiology and Biostatistics, Western University London, ON, Canada; Department of Medicine (Geriatrics), Dalhousie University, Halifax, NS, Canada; Public Health Ontario, Toronto, Ontario, Canada; ICES, Toronto, Ontario, Canada

## Abstract

**Background:** As COVID-19 becomes established as an endemic disease with widespread population immunity, there is uncertainty about the economic benefit of ongoing COVID-19 vaccination programs. We assessed the cost-effectiveness of a COVID-19 vaccination program similar to current Canadian recommendations, modelled as annual vaccination for people aged less than 65 years with chronic medical conditions and biannual vaccination for adults aged 65 years and older.

**Methods:** Using a static individual-based model of medically attended COVID-19 in a population of 1 million people, we estimated costs (in 2023 Canadian dollars), quality-adjusted life years (QALYs), and incremental cost-effectiveness ratios (ICERs). We used health system and societal perspectives and a 1.5% discount rate. Parameters were based on recent COVID-19 epidemiology, vaccine characteristics, and costs.

**Results:** Between July 2024 and September 2025, a program similar to current Canadian recommendations was estimated to avert 3.1% (95% credible interval (CrI): 3.0 to 3.2%) of outpatient cases, 8.8% (95% CrI: 7.3 to 10.4%) of inpatient cases, 3.6% (95% CrI: 2.8 to 4.3%) of PCC cases, and 9.4% (95% CrI: 5.6 to 13.8%) of deaths compared to no vaccination. The number needed to vaccinate to prevent one hospitalization and one death was 1,121 (95% CrI: 941 to 1,357) and 8,656 (95% CrI: 5,848 to 14,915), respectively. For the health system perspective, the program would cost an additional $4.695 million but result in 221.17 QALYs gained, leading to an ICER of $21,227 per QALY compared to no vaccination. Vaccine price influenced cost-effectiveness, with higher prices reducing the likelihood the program met common cost-effectiveness thresholds.

**Conclusions:** A program similar to current COVID-19 recommendations in Canada is likely effective and cost-effective compared to no vaccination. However, unlike some other research studies, alternate vaccination strategies that may offer better value for money were not evaluated.

## INTRODUCTION

The value of vaccination during the COVID-19 pandemic has been well documented (1–4). Between December 2020 and March 2022, Canada’s COVID-19 vaccination program was estimated to result in tens to hundreds of billions of dollars in monetary benefit due to prevention of premature mortality and reduced illness and disability (1). COVID-19 has transitioned from a pandemic to an endemic phase; however, it has not established the clear fall/winter-time seasonal patterns that are observed with some other respiratory viruses, such as influenza and respiratory syncytial virus (RSV) (5–8). Given the less predictable timing of increases in COVID-19 case occurrence, relatively short observed duration of protection following vaccination, and high levels of population level immunity due to prior infection and/or vaccination, the impact and value for money for ongoing COVID-19 vaccination programs is less clear than during the pandemic phase.

Canada’s National Advisory Committee on Immunization (NACI) has been issuing guidance on COVID-19 vaccination for the fall and spring for the past several years. For the fall of 2024, NACI has strongly recommended COVID-19 vaccination for people aged 65 years and older and people aged 6 months to 64 years who are at increased risk of SARS-CoV-2 infection or severe COVID-19 disease (9). For other groups 6 months to 64 years of age who are at average risk of severe disease, NACI has recommended a discretionary annual dose (10).

Advice for spring 2025 has not yet been issued, but NACI has typically had a discretionary recommendation for an additional vaccine dose received at a 6-month interval (with a minimum interval of 3 months) for those aged 65 years and older, adult residents of long-term care homes and other congregate living settings for seniors, and those aged 6 months to 64 years who are moderately to severely immunocompromised (10).

Prior to 2025, Canadian provinces and territories accessed COVID-19 vaccines procured through federal pandemic investments, with vaccine costs covered by the federal government. Starting in 2025, provinces and territories will switch to regular procurement pathways and will assume the cost of purchasing COVID-19 vaccines for their populations (9). As increasing scrutiny is given to the costs and benefits of a COVID-19 vaccination program in the current epidemiological context, we sought to evaluate the cost-utility of a vaccination strategy similar to recommendations in Canada. This evaluation was conducted as a part of an economic evidence package used to inform NACI recommendations for the use of COVID-19 vaccines in Canada from 2025 to summer of 2026.

## METHODS

### Model overview

We conducted a model-based cost-utility analysis of COVID-19 vaccination programs in Canada, comparing a strategy similar to the key features of Canada’s currently recommended program (biannual vaccination for adults aged 65 years and older and annual vaccination for the population aged less than 65 years with one or more chronic medical conditions) to a “no vaccination” strategy (i.e., no COVID-19 vaccination occurring in the population during the model time period of July 2024 to September 2025). For brevity, we refer to the modelled vaccination strategy as the “current recommendations” but note that it is simplification of the actual recommendations. We conducted extensive scenario analyses to examine how alternate assumptions about vaccine costs, program timing, and epidemiology would impact results.

The model used monthly time steps and included the time period from July 2024 to September 2025, to evaluate an annual vaccination program. The model start time allowed for the exploration of an earlier program start time in scenario analyses. We measured costs in 2023 Canadian dollars; where necessary, costs were adjusted using the Canadian Consumer Price Index (11). We measured health outcomes as quality-adjusted life years (QALYs). We used an annual discount rate of 1.5% for long-term costs and outcomes and assessed cost-effectiveness from the health system and societal perspectives (12). We used R to construct the model and conduct the analysis (13). The study followed the ISPOR Consolidated Health Economic Evaluation Reporting Standards (CHEERS) 2022 reporting guidance (14).

### Model structure

We adapted a static individual-based cost-effectiveness model of medically attended COVID-19 from a previously described model of RSV disease (15). The model followed a closed population of one million people stratified by age group and medical-risk status (**Figure 1**) (16–18). To evaluate current vaccination recommendations for people at higher risk (HR) for experiencing severe outcomes following a SARS-CoV-2 infection, we used data on the proportion of the population in Canada with one or more chronic medical conditions (CMCs) (17, 18). The remaining proportion of the population without CMCs was characterized as average risk (AR).

**Figure 1.**
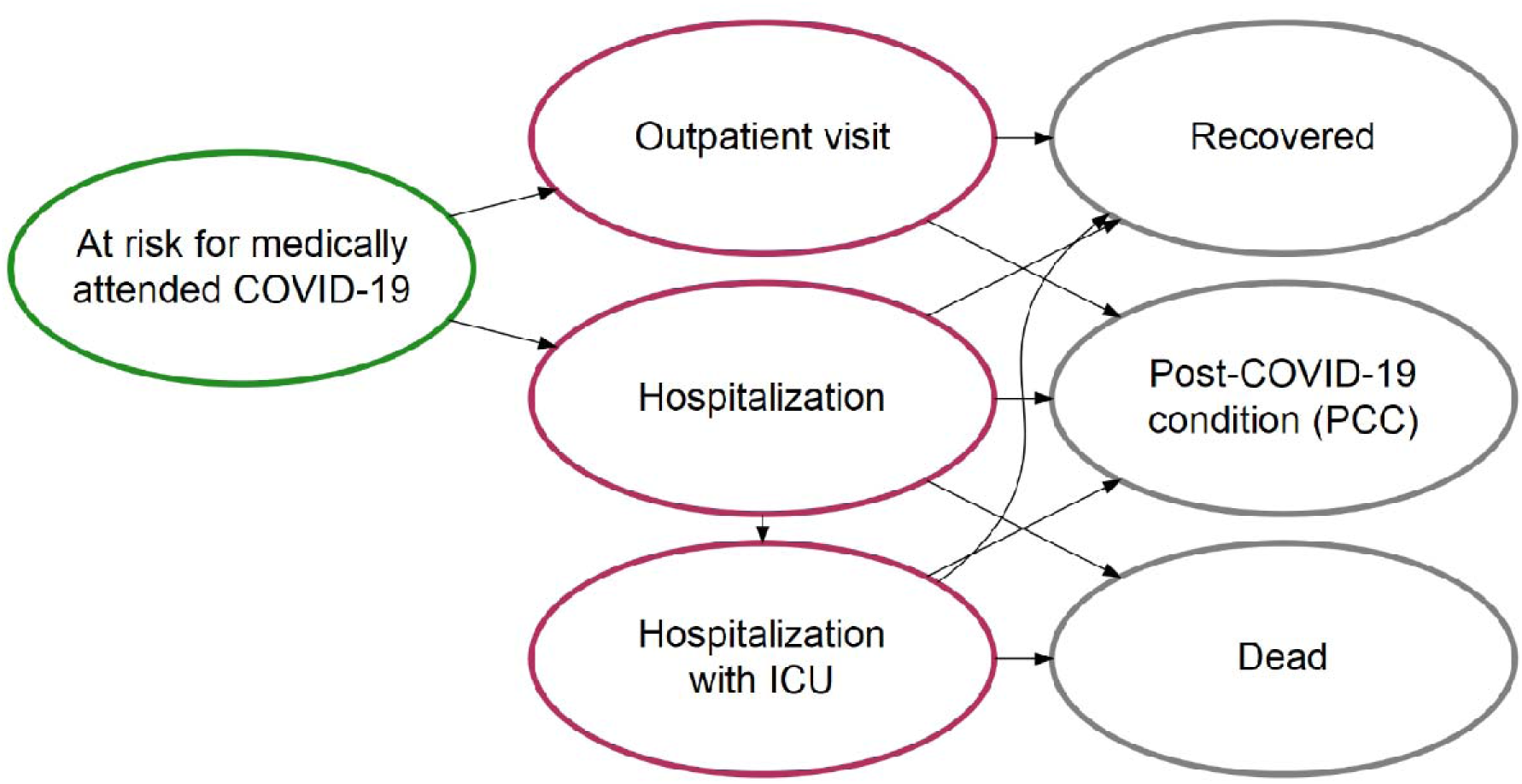
Overview of health states included in the cost-utility model. Health states and transitions between states are shown. Transitions from all health states to death were also possible due to background mortality but are not shown in the schematic. The model was stratified by age group and medical risk status, defined as the presence or absence of at least one chronic medical condition, to allow for age and medical risk dependent transition probabilities.

### Model parameters

Parameters describing COVID-19 epidemiology, vaccine characteristics, health utilities, and costs are provided in **Table 1** and **Table 2**. We used published studies, with a preference for Canadian data, when available. When data were unavailable, we used assumption. Where possible, we used data stratified by age group and/or medical-risk status. For parameters with ranges specified in the tables, values were sampled from gamma distributions for costs and beta distributions for probabilities and utilities. Additional details for key parameters and assumptions are provided below.

**Table 1.** Model parameters: population, SARS-CoV-2 epidemiology, and vaccine characteristics.

| Parameter | Base | Range | Reference |
| --- | --- | --- | --- |
| Population distribution (%) |  |  |  |
| Less than 5 years | 4.7 | -- | Statistics Canada (16) |
| 5-19 years | 13.9 | -- |  |
| 20-49 years | 42.7 | -- |  |
| 50-64 years | 19.1 | -- |  |
| 65-74 years | 11.1 | -- |  |
| 75+ years | 8.6 | -- |  |
| % of population with one or more chronic medical conditions (CMCs) |  |  |  |
| Less than 5 years | 14.4 | -- | CPCSSN (17); Statistics Canada (40); Assumption |
| 5-19 years | 21.0 | -- |  |
| 20-49 years | 24.9 | -- |  |
| 50-64 years | 45.6 | -- |  |
| 65-74 years | 60.1 | -- |  |
| 75+ years | 72.1 | -- |  |
| Monthly % of annual COVID-19 cases: hospitalization data |  |  |  |
| July | 3.7 | -- | Government of Canada (20) |
| August | 3.7 | -- |  |
| September | 12.5 | -- |  |
| October | 15.2 | -- |  |
| November | 16.0 | -- |  |
| December | 19.0 | -- |  |
| January | 10.4 | -- |  |
| February | 6.1 | -- |  |
| March | 4.7 | -- |  |
| April | 3.0 | -- |  |
| May | 3.7 | -- |  |
| June | 1.9 | -- |  |
| Monthly % of annual COVID-19 cases: Respiratory Virus Detection Surveillance System data |  |  |  |
| July | 5.1 | -- | RVDSS (41) |
| August | 8.4 | -- |  |
| September | 12.4 | -- |  |
| October | 13.4 | -- |  |
| November | 13.2 | -- |  |
| December | 12.4 | -- |  |
| January | 8.7 | -- |  |
| February | 5.5 | -- |  |
| March | 4.2 | -- |  |
| April | 3.7 | -- |  |
| May | 5.7 | -- |  |
| June | 7.4 | -- |  |
| Monthly % of annual COVID-19 cases: model projection |  |  |  |
| July | 2.9 | -- | Transmission model (19) |
| August | 5.3 | -- |  |
| September | 8.7 | -- |  |
| October | 14.2 | -- |  |
| November | 19.6 | -- |  |
| December | 20.8 | -- |  |
| January | 10.8 | -- |  |
| February | 5.9 | -- |  |
| March | 4.2 | -- |  |
| April | 2.9 | -- |  |
| May | 2.5 | -- |  |
| June | 2.1 | -- |  |
| Annual probability of medically attended COVID-19 treated in an outpatient setting |  |  |  |
| 0-4 years | 0.106 | -- | Transmission model (19) |
| 5-19 years | 0.075 | -- |  |
| 20-49 years | 0.075 | -- |  |
| 50-64 years | 0.106 | -- |  |
| 65+ years | 0.120 | -- |  |
| Relative risk of medically attended outpatient care in people with one or more CMCs compared to people without a CMC |  |  |  |
| All ages | 1.5 | -- | Assumption |
| Annual incidence of COVID-19-attributable hospitalization per 100,000 population |  |  |  |
| 0-4 years | 33.7 | -- | Transmission model (19) |
| 5-19 years | 11.6 | -- |  |
| 20-49 years | 20.6 | -- |  |
| 50-64 years | 64.0 | -- |  |
| 65+ years | 506.9 | -- |  |
| % of patients hospitalized with confirmed COVID-19 with one or more CMCs |  |  |  |
| 0-4 years | 49.3 | -- | COVID-NET (42) |
| 5-19 years | 53.7 | -- |  |
| 20-49 years | 82.2 | -- |  |
| 50-64 years | 97.3 | -- |  |
| 65+ years | 98.9 | -- |  |
| % of patients hospitalized with COVID-19 requiring ICU admission |  |  |  |
| All ages | 13.2 | 12.7-13.6 | CIHI, 2024 (43) |
| COVID-19 mortality per hospitalization (%) |  |  |  |
| 0-4 years | 2.9 | -- | Transmission model (19);<br>Centers for Disease Control<br>and Prevention (44);<br>Assumption |
| 5-19 years | 2.9 | -- |  |
| 20-49 years | 3.3 | -- |  |
| 50-64 years | 9.5 | -- |  |
| 65-74 years | 13.7 | -- |  |
| 75+ years | 17.9 | -- |  |
| All cause mortality rate per year, per 1,000 population |  |  |  |
| All ages | Age-specific<br>rates | -- | Statistics Canada (45) |
| Outpatient COVID-19 duration (days) |  |  |  |
| All ages | 5 | 1-10 | Centers for Disease Control<br>and Prevention (46);<br>Government of Ontario (47) |
| Length of stay in hospital (days) |  |  |  |
| All ages | 24 | 16.2-48.6 | CIHI, 2024 (43) |
| Length of stay in ICU (days) |  |  |  |
| All ages | 9.4 | 2.6-15.9 | CIHI, 2024 (43) |
| % of patients with medically attended COVID-19 prescribed an antiviral |  |  |  |
| 20+ years | 3.1 | 2.5-3.7* | Kuang et al. 2023 (48) |
| Incremental vaccine effectiveness for COVID-19 infection requiring outpatient medical care, by<br>time since vaccination (%) |  |  |  |
| First 2 months | 50 |  | ACIP (28) |
| 3 to 4 months | 32 |  |  |
| 5 to 6 months | 1 |  |  |
| Incremental vaccine effectiveness for COVID-19 infection requiring hospitalization, by time since<br>vaccination (%) |  |  |  |
| First 2 months | 49 |  | ACIP (28) |
| 3 to 4 months | 43 |  |  |
| 5 to 6 months | 14 |  |  |
| Vaccine effectiveness weight for people with one or more CMCs |  |  |  |
| All ages | 0.8 |  | Assumption |
| Vaccination coverage (%) – first (fall) dose |  |  |  |
| 0-4 years | 5.7 |  | Public Health Agency of<br>Canada (27) |
| 5-19 years | 6.3 |  |  |
| 20-49 years | 9.1 |  |  |
| 50-64 years | 23.8 |  |  |
| 65+ years | 52.7 |  |  |
| Vaccination coverage (%) – second (spring) dose |  |  |  |
| 65+ years | 11 |  | Public Health Agency of<br>Canada (27) |
| Time to receive vaccine (hours), including travel and waiting time |  |  |  |
| All ages | 2 | 1.5-2.5 | Canada Health Infoway (49); Ray et al. 2015 (50) |
| Vaccine wastage rate (%) |  |  |  |
| All ages | 10 (10-30) | -- | Office of the Auditor General of Ontario (51); assumption |
| Adverse events following immunization, per 100,000 doses administered |  |  |  |
| Serious adverse event | 1.1 | -- | Public Health Agency of Canada (52) |
| Duration of adverse event following immunization (days) |  |  |  |
| Serious adverse event | 2 | 1-4 | Lee et al. 2009 (53) |
| Probability of developing post-COVID-19 condition (PCC) among outpatient cases |  |  |  |
| 0-4 years | 0.005 | 0.003-0.006 | ACIP (21); Assumption |
| 5-19 years | 0.007 | 0.005-0.009 |  |
| 20+ years | 0.023 | 0.017-0.029 |  |
| Probability of developing PCC among hospitalized cases |  |  |  |
| 0-4 years | 0.008 | 0.003-0.013 | ACIP (21); Assumption |
| 5-19 years | 0.012 | 0.005-0.020 |  |
| 20+ years | 0.040 | 0.017-0.064 |  |
| Duration of PCC (days) |  |  |  |
| All ages | 365 | -- | CCAHS (54) |
| % of PCC cases that miss school or work | 16 | -- |  |
| Duration of school or work missed due to PCC | 23.3 | -- |  |

\*Range defined as $\pm 20\%$ of the base value

**Table 2.** Model parameters: costs and utilities.

| Parameter | Base | Range | Reference |
| --- | --- | --- | --- |
| Vaccination cost per dose (\$) | | | |
| Vaccine | 43 | 43-107 | Centers for Disease Control and Prevention (55); Mulcahy et al. 2024 (56); assumption |
| Administration | 18 | 13-22 | O'Reilly et al. 2017 (57) |
| Transportation and storage | 0.64 | -- | Sah et al. 2022 (2) |
| Attributable medical costs per person with COVID-19 not requiring hospitalization (\$) | | | |
| All ages | 295 | 207-383 | Sander et al. 2024 (26) |
| Attributable medical costs per person hospitalized with COVID-19 (\$) | | | |
| Hospitalization without ICU admission | 30,147 | 28,760-31,534 | Sander et al. 2024 (26) |
| Hospitalization with ICU admission | 105,677 | 100,467-110,888 |  |
| Direct medical costs for severe adverse event following vaccination (\$) | | | |
| <65 years | 62 | 48-82 | Sander et al. 2010 (58); CIHI (59); Lee et al. 2009 (53) |
| 65+ years | 66 | 51-87 |  |
| Transportation costs (\$) | | | |
| Cost of two-way travel to vaccination or outpatient care (out of pocket) | 14 | 11-16* | Canada Health Infoway (49, 60) |
| Cost of travel to inpatient care | 417 | 210-623 | NACI (60) |
| Co-pay per prescription (\$) | | | |
| 20+ years | 10 | 5-12 | University of Western Ontario (61); The Commonwealth Fund (62); assumption |
| Out-of-pocket medication costs (\$) | | | |
| Over-the-counter medication costs for medically attended case | 12 | 3-20 | Federici et al. 2018 (63) |
| Prescription medication costs for severe adverse event following vaccination (<65 years) | 4.19 | 3.14-5.24 | Lee et al. 2009 (53) |
| Antiviral costs (20-64 years) | 1,289 | 1,031-1,547* | CADTH, 2024 (64) |
| Caregiver workdays lost |  |  |  |
| % reduction in productivity | 33 | -- | Keita Fakeye et al. 2023 (65) |
| Labour force participation (%) |  |  |  |
| 5-19 years | 16.9 | -- | Statistics Canada (66) |
| 20-49 years | 87.1 | -- |  |
| 50-64 years | 74.6 | -- |  |
| 65-74 years | 18.7 | -- |  |
| 75+ years | 8.0 | -- |  |
| Caregiver | 88.8 | -- |  |
| Average employment income (\$) | | | |
| 5-19 years | 20,600 | -- | Statistics Canada (67) |
| 20-49 years | 60,598 | -- |  |
| 50-64 years | 70,727 | -- |  |
| 65+ years | 50,058 | -- |  |
| Caregiver (age 25-54 years) | 70,452 | -- |  |
| Background health utility |  |  |  |
| 0-4 years | 0.950 | -- | Assumption |
| 5-19 years | 0.917 | -- | Molina et al. 2023 (22) |
| 20-49 years | 0.880 | -- |  |
| 50-64 years | 0.845 | -- | Yan et al. 2023 (23) |
| 65-74 years | 0.867 | -- |  |
| 75+ years | 0.861 | -- |  |
| QALY loss, outpatient care |  |  |  |
| 0-4 years | 0.00575 | 0.00301-0.0085 | Soare 2023 (24) |
| 0-19 years | 0.00561 | 0.00287- 0.00835 |  |
| 20+ years | 0.0045 | 0.0018-0.0074 |  |
| QALY loss, hospitalization without ICU admission |  |  |  |
| 0-19 years | 0.01875 | 0.0052-0.0324 | Soare 2023 (24) |
| 20+ years | 0.0174 | 0.0038-0.031 |  |
| QALY loss, hospitalization with ICU admission |  |  |  |
| 0-19 years | 0.08341 | 0.0592-0.111 | Mercon 2023 (25) |
| 20+ years | 0.0394 | 0.0231-0.0583 |  |
| QALY loss, PCC following outpatient care |  |  |  |
| 0-4 years | 0.1206 | 0.0899-0.1568 | Assumption |
| 5-20 years | 0.1126 | 0.0830-0.1481 | Mercon 2023 (25) |
| 20+ years | 0.0608 | 0.038-0.0912 |  |
| QALY loss, PCC following infection treated in hospital |  |  |  |
| 0-4 years | 0.1412 | 0.1053-0.1836 | Assumption |
| 5-20 years | 0.1319 | 0.09719-0.1734 | Mercon 2023 (25) |
| 20+ years | 0.0712 | 0.0445-0.1068 |  |
| QALY loss, death |  |  |  |
| 0-4 years | 41.38 | -- | Statistics Canada (45, 68) |
| 5-19 years | 38.15 | -- |  |
| 20-49 years | 29.58 | -- |  |
| 50-64 years | 19.07 | -- |  |
| 65-74 years | 13.14 | -- |  |
| 75+ years | 7.35 | -- |  |
| Quality-adjusted life days lost, adverse event following vaccination |  |  |  |
| Serious adverse event | 1.0 | 0.75-1.25 | Sandmann et al. 2021 (69) |

\*Range defined as $\pm 20\%$ of the base value

### COVID-19 epidemiology

To estimate annual cumulative incidence of symptomatic and hospitalized COVID-19 cases in the absence of any COVID-19 vaccination during the model time period (i.e., the no vaccination counterfactual) we used a separate age-stratified dynamic compartmental transmission model. The model accounts for boosting and waning of immunity in the context of infection and vaccination (19) and was calibrated to COVID-19 hospital occupancy (from January 2022 to April 2024) and seroprevalence data (from January 2022 to December 2023). We projected COVID-19 incidence between July 2024 and September 2025 without any vaccination occurring in the population. We then used these annualized incidence estimates for the no vaccination counterfactual scenario as inputs in our static cost-effectiveness model to estimate the impact of vaccination for preventing medically attended COVID-19 cases.

For the base-case analysis, we assumed that the proportion of annual COVID-19 cases occurring each month followed the distribution of hospitalized cases reported from July 2023 to June 2024 (20), with alternate case distributions used in scenario analyses (**Figure 2A**). We only included costs and QALYs associated with medically attended COVID-19, which was defined as requiring outpatient (i.e., health care provider or emergency department visit) or inpatient (i.e., hospital admission, with or without intensive care unit (ICU) admission) care. We assumed that a proportion of people with medically attended COVID-19 developed post-COVID condition (PCC), with risk of PCC dependent on severity of disease (21). We also included costs and QALY losses associated with COVID-19 mortality.

**Figure 2.**
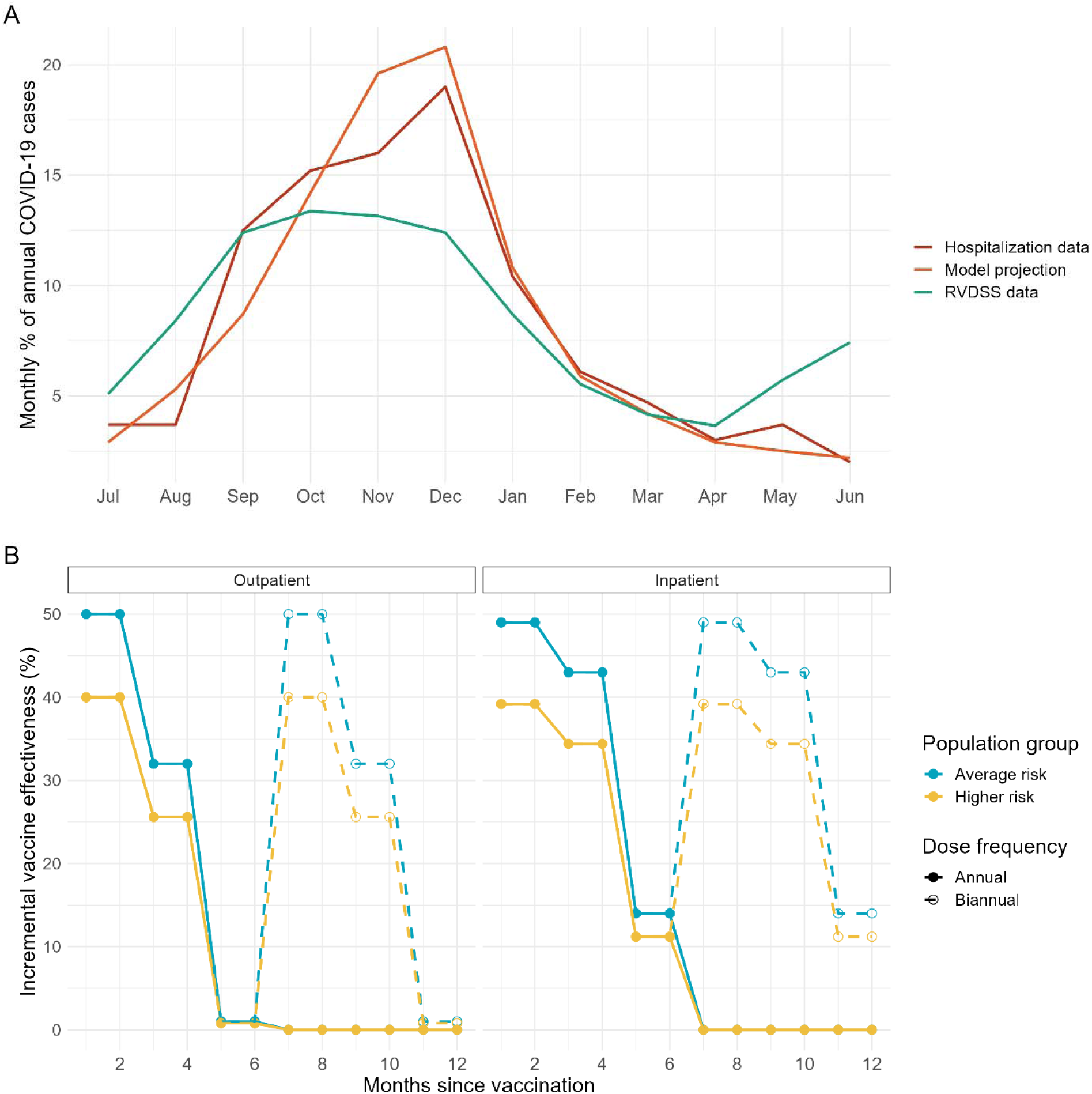
Model assumptions for monthly distribution of COVID-19 cases and vaccine effectiveness. (A) The assumed distribution of COVID-19 cases over the course of the year. The base-case analysis used the distribution based on hospitalization data and distributions based on Respiratory Virus Detection Surveillance System (RVDSS) data (“flatter curve” scenario) and projections from a transmission model (“larger winter wave” scenario) were evaluated in scenario analyses. (B) Incremental vaccine effectiveness (VE) was assumed to vary by outcome (medically attended COVID-19 treated in an outpatient or inpatient setting), risk status (lower for higher risk than average risk), and time since vaccination. If receiving two COVID-19 vaccines a year, receipt of the second dose was assumed to return VE to initial values and result in similar waning over time. People with one or more chronic medical conditions were considered higher risk and people without chronic medical conditions were considered average risk. The same vaccine effectiveness for the high risk population group as the average risk group was evaluated in a scenario analysis.

### Utilities

We used Canadian age-specific utilities based on EQ-5D-5L index scores to calculate QALY losses due to COVID-19 mortality (22, 23). QALY losses due to other COVID-related health outcomes were derived from published studies (24, 25) and assumptions.

### Costs

We used health care costs attributable to diagnosed COVID-19 from a population-based matched cohort study in Ontario, Canada, which included both acute care and post-acute care costs for an approximately one-year period following initial diagnosis for people treated in outpatient and inpatient settings (26). Because post-acute care costs were assumed to include costs associated with PCC, additional health care costs for PCC were not included.

As there were no established Canadian vaccine list prices at the time of the analysis, we assumed a price of 40% ($43) of the US Centers for Disease Control and Prevention (CDC) public list price of $107 per dose. This estimate was based on unpublished Public Health Agency of Canada data suggesting that Canadian negotiated vaccine prices are typically 30-50% of US public list prices. We also evaluated prices of 75% ($80) and 100% ($107) of the US CDC list price in scenario analyses. Other vaccination costs included vaccine administration, vaccine wastage, and adverse events following immunization (AEFIs).

For the societal perspective, we included the following costs: patient productivity loss due to COVID-attributable disease and death, vaccination and AEFIs, caregiver productivity loss, and out-of-pocket medical costs. We estimated productivity loss using the human capital approach and used the friction cost approach in a sensitivity analysis. We used age-specific labour force participation rates and average employment income (12, 15).

### Vaccination

In the base-case analysis, annual vaccination occurred in October and November. For the population aged 65 years and older who received a second dose, it was administered six months after receipt of the first dose (i.e., April or May). The proportion of the population vaccinated varied by age and was based on Canadian estimates of uptake in the spring 2023 and fall/winter 2023-2024 vaccination campaigns. For example, uptake was 53% in the fall and 11% in the spring for those aged 65 years and older (27). For those aged less than 65 years, who only received one dose, coverage increased with increasing age and ranged from 5.7% in the under 5 year age group to 23.8% in the 50 to 64 year age group.

We used US observational data for the 2023-2024 season (28) to inform estimates of incremental vaccine effectiveness (VE) and waning. We assumed VE against more severe disease (i.e., COVID-19 requiring hospitalization) waned more slowly than VE against less severe disease (i.e. COVID-19 treated in an outpatient setting); however, incremental VE was assumed to reach 0 for protection against all outcomes by the seventh month following vaccination (**Figure 2B**). We also assumed lower incremental VE for people at higher risk of COVID-19 compared to those at average risk. Vaccination lowered the risk of PCC by decreasing the overall likelihood of SARS-CoV-2 infection, and additional VE for preventing PCC in people with medically attended COVID-19 was not included.

### Analysis

We calculated QALYs and costs associated with the modelled health outcomes for the two strategies (i.e., a vaccination program similar to current recommendations versus no further vaccination from July 2024 onward) and calculated the ICER. QALYs, costs, and the ICER are the mean of 2,000 model simulations, each based on a draw from parameter distributions. The number of simulations was chosen to ensure sufficient sampling of the probability distributions. For the base case analysis, we also calculated net monetary and net health benefit, which represent the value of the vaccination strategy in terms of monetary value and QALYs, respectively (12). Since Canada does not have a cost-effectiveness threshold, we applied the commonly used $50,000 per QALY threshold (29) to the net benefit calculations.

We calculated outcomes averted compared to no vaccination and number needed to vaccinate to avert an outpatient case (i.e., health care provider or emergency department visit), inpatient case, PCC case, or death. Number needed to vaccinate was calculated as number of doses administered divided by outcomes averted. Summary results for health outcomes represent medians and 95% credible intervals (Crl) from 2,000 model simulations.

### Sensitivity and scenario analyses

We conducted a probabilistic sensitivity analysis for the base-case analysis to estimate the probability that each vaccination strategy was cost-effective at varying cost-effectiveness thresholds. We also conducted scenario analyses focusing on alternate assumptions relating to vaccine wastage, vaccine prices, COVID-19 incidence, monthly distribution of COVID-19 cases, vaccination coverage, and vaccination program timing relative to the period of highest disease activity. Details of the scenarios are provided in **Table 3**. For the societal perspective, we repeated our analysis using the friction cost approach. The friction cost approach differs from the human capital approach by assuming that after a “friction period” (3 months in our analysis), workers who have left the workforce will be replaced by currently unemployed workers (12).

**Table 3.** Summary of scenario analyses and incremental cost-effectiveness ratios (ICERs) for the current vaccination strategy compared to no vaccination.

| Scenario | Scenario assumptions* | ICER (\$ per QALY) for current vaccination strategy** compared to no vaccination | |
| --- | --- | --- | --- |
|  |  | Health system perspective | Societal perspective |
| Base case | • \$43 per dose of vaccine (40% of CDC list price)<br>• 10% vaccine wastage<br>• First dose vaccination coverage increases with age: 5.7% (<5 years); 6.3% (5 to 19 years); 9.1% (20 to 49 years); 23.8% (50 to 64 years); 52.7% (65 years and older)<br>• VE for HR population is 0.8-fold the VE for AR population<br>• Median estimate of COVID incidence from transmission model<br>• Monthly distribution of cases estimated from 2023-2024 hospitalization data<br>• Annual dose vaccine protection starts in October/November<br>• Second dose protection starts in April/May for the population aged 65 and older receiving a second dose | 21,227 | 25,867 |
| Higher vaccine price (\$80 per dose) | • \$80 per dose of vaccine (75% of CDC list price) | 51,388 | 56,029 |
| Higher vaccine price (\$107 per dose) | • \$107 per dose of vaccine (100% of CDC list price) | 73,398 | 78,039 |
| Higher vaccine wastage (20%) | • 20% vaccine wastage | 24,413 | 29,053 |
| Higher vaccine wastage (30%) | 30% vaccine wastage | 27,600 | 32,240 |
| Higher vaccination uptake for younger age groups | 52.7% first dose vaccination coverage for all age groups | 41,291 | 54,406 |
| Equal VE for HR and AR groups | VE for HR population is the same as the VE for AR population | 11,964 | 13,382 |
| Higher COVID-19 incidence | COVID-19 incidence assumed to be 2-times the base-case value | 21,409 | 26,043 |
| Alternate seasonality (flatter curve) | Monthly distribution of cases estimated from 2023-2024 Respiratory Virus Detection Surveillance System data | 30,822 | 39,389 |
| Alternate seasonality (larger winter wave) | Monthly distribution of cases estimated from transmission model | 17,725 | 20,882 |
| Earlier program start | Annual dose vaccine protection starts in August/September | 20,240 | 25,197 |
| Earlier program start and 4-month interval between doses | Annual dose vaccine protection starts in August/September<br>Second dose protection starts in December/January for the population aged 65 and older receiving a second dose | 18,535 | 22,943 |

\*For each scenario except the base case, any changes from the base case are indicated. All relevant base-case scenario assumptions are provided for reference.
\*\*Current strategy modelled as 2 doses for all adults aged 65 years and older and an annual dose for the population at higher risk of COVID-19 aged less than 65 years.
Note: AR = average risk (no chronic medical conditions); HR = higher risk (one or more chronic medical conditions).

## RESULTS

### Impact of a vaccination strategy with key features of the currently recommended vaccination program

For the no vaccination strategy, we estimated 8,665 (95% CrI: 8,615 to 8,715) outpatient cases, 134 (95% CrI: 126 to 140) inpatient cases, 177 (95% CrI: 137 to 223) PCC cases, and 16 (95% CrI: 14 to 18 ) deaths per 100,000 person-years for the time period from July 2024 to September 2025. Compared to the no vaccination strategy, we estimated that the currently recommended COVID-19 vaccination program would avert 3.1% (95% CrI: 3.0 to 3.2%) of outpatient cases, 8.8% (95% CrI: 7.3 to 10.4%) of inpatient cases, 3.6% (95% CrI: 2.8 to 4.3%) of PCC cases, and 9.4% (95% CrI: 5.6 to 13.8%) of deaths, assuming the same vaccine coverage as estimated for 2023/2024.

Number needed to vaccinate was: 50 (95% CrI: 48 to 51) to prevent one outpatient case, 1,121 (95% CrI: 941 to 1,357) to prevent one hospitalization, 2,079 (95% CrI: 1,544 to 2,969) to prevent one PCC case, and 8,656 (95% CrI: 5,848 to 14,915) to prevent one death. When estimates of number needed to vaccinate were stratified by the age groups included in the current recommendations (under 65 years or 65 years and older), we observed similar number needed to vaccinate values across groups for prevention of outpatient and PCC cases.

However, number needed to vaccinate for prevention of one hospitalization was higher for the population aged less than 65 years (4,250, 95% CrI: 2,421 to 11,297) than for those aged 65 years and older (1,088, 95% CrI: 921 to 1,310). Due to the low number of projected deaths in the under 65 year age group, number needed to vaccinate to prevent one death was not estimable for this age group, but was 7,228 (95% CrI: 4,812 to 12,939) for the population aged 65 years and older.

### Base-case analysis

Compared to no vaccination, a program based on the key features of the current program recommendations, including current uptake levels, would result in $4.695 million in additional costs and 221.17 QALYs gained in a population cohort of 1 million people, resulting in an ICER of $21,227 per QALY for the health system perspective (**Table 4**). For the societal perspective, incremental costs were higher ($5.721 million), resulting in an ICER of $25,867. Using a cost-effectiveness threshold of $50,000 per QALY, the current vaccination strategy compared to no vaccination resulted in a net monetary benefit of $6.34 million and a net health benefit of 127.28 QALYs in a population of 1 million people for the health system perspective. For the societal perspective, net monetary benefit was $5.34 million and net health benefit was 106.75 QALYs. Positive net benefit indicates greater benefits than costs and an increase in population health associated with the currently recommended vaccination strategy compared to no vaccination.

**Table 4.** Costs, QALYs, and ICERs for the base-case analysis for the health system and societal perspectives.

| | Costs (\$) | | | Effects (QALYs lost) | Incremental costs (\$) | | | Incremental effect (QALYs gained) | ICER (\$ per QALY gained) |
| --- | --- | --- | --- | --- | --- | --- | --- | --- | --- |
|  | Medically attended COVID-19 | Vaccination program | Total |  | Medically attended COVID-19 | Vaccination program | Total |  |  |
| Health system perspective |  |  |  |  |  |  |  |  |  |
| No vaccination | 90,643,206 | -- | 90,643,206 | 2,905.988 | -- | -- | -- | -- | -- |
| Current recommendations | 84,529,887 | 10,808,083 | 95,337,970 | 2,684.814 | - 6,113,319 | 10,808,083 | 4,694,764 | 221.1735 | 21,227 |
| Societal perspective |  |  |  |  |  |  |  |  |  |
| No vaccination | 196,120,300 | -- | 196,120,300 | 2,905.988 | -- | -- | -- | -- | -- |
| Current recommendations | 186,941,423 | 14,899,930 | 201,841,353 | 2,684.814 | - 9,178,877 | 14,899,930 | 5,721,053 | 221.1735 | 25,867 |

Probabilistic sensitivity analysis showed that the currently recommended program had a high likelihood of being cost-effective compared to no vaccination for commonly used cost-effectiveness thresholds (**Figure 3**). Using a $50,000 per QALY threshold, probabilities were 0.993 (health system perspective) and 0.967 (societal perspective).

**Figure 3.**
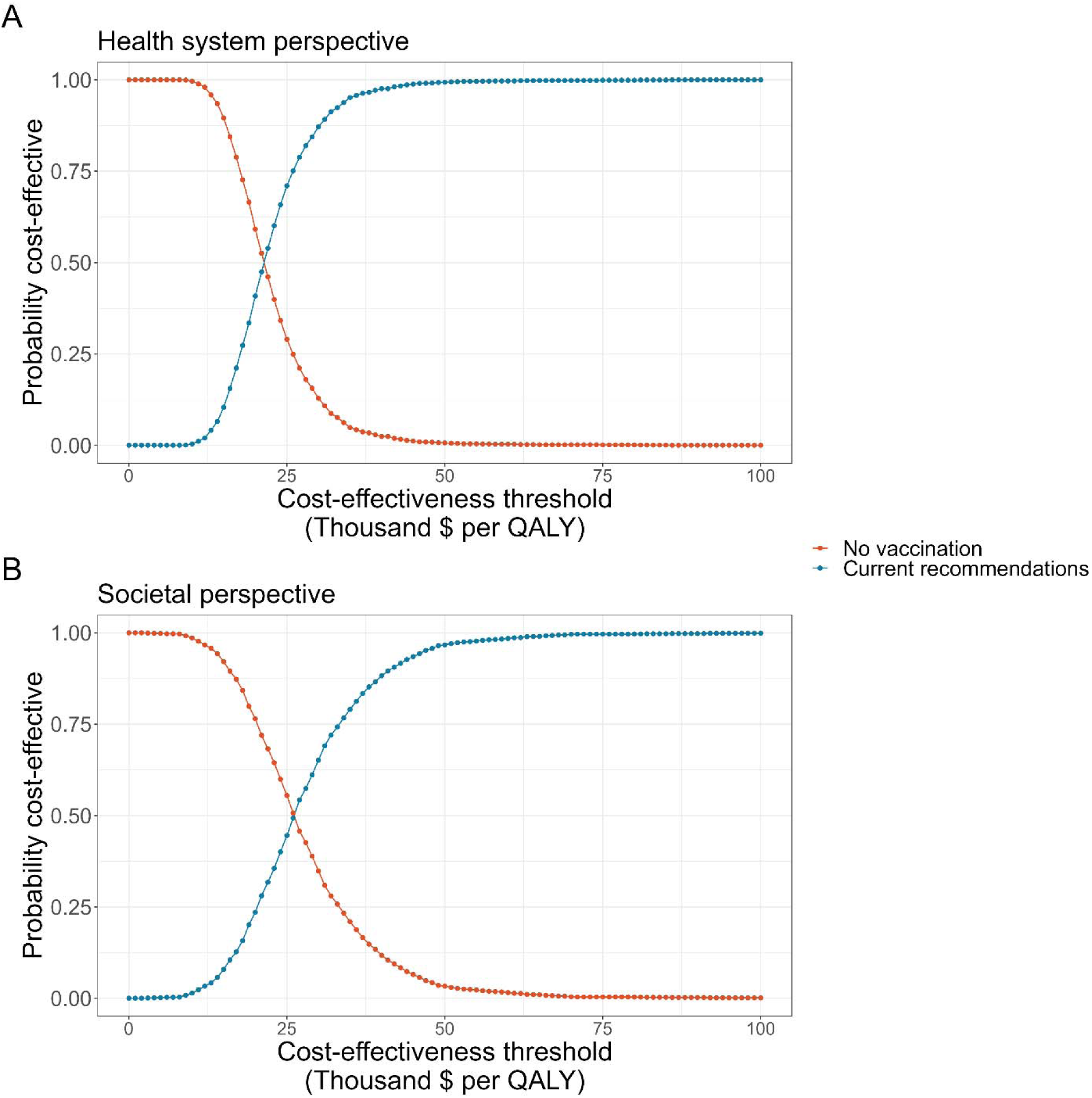
Proportion of simulations that each strategy was cost-effective at a given cost-effectiveness threshold. Cost-effectiveness acceptability curves are based on probabilistic sensitivity analysis for 2,000 model simulations for the (A) health system and (B) societal perspectives. Points show the proportion of samples for which each strategy was identified as cost-effective at given cost-effectiveness threshold.

### Scenario analyses

ICERs for the strategy similar to current recommendations compared to no vaccination were lower than the base case for the following scenarios using alternate assumptions: no reduction in VE for people at high risk of COVID compared to those at average risk; a higher concentration of COVID-19 cases occurring in the winter months, or an earlier annual vaccination program start date (with or without a shorter interval between doses for those receiving two doses) (**Table 3**). ICERs in these scenarios ranged from $11,964 to $20,240 per QALY for the health system perspective and $13,382 to $25,197 per QALY for the societal perspective. ICERs were higher than base-case values for all other scenarios considered but remained less than $50,000 per QALY for the health system perspective, with the exception of a higher assumed vaccine price. For a vaccine price of $80 per dose, the ICER was $51,388 per QALY and for a vaccine price of $107 per dose, the ICER was $73,398 per QALY for the current recommendations compared to no vaccination for the health system perspective. For the societal perspective, ICERs for the current recommendations were $56,029 per QALY and $78,039 per QALY compared to no vaccination when vaccine price was $80 or $107 per dose, respectively, and $54,406 per QALY when higher vaccination uptake was assumed for the under 65 year old population at higher risk for COVID-19.

Using the friction cost approach to quantify lost productivity related to paid work for the societal perspective, we found that compared to no vaccination, a program based on current recommendations would result in higher incremental costs ($6.756 million) than when using the human capital approach ($5.721 million). Consequently, the ICER for the current program was also higher, at $30,545 per QALY, than was estimated using the human capital approach ($25,867 per QALY).

## DISCUSSION

Our economic evaluation of a COVID-19 vaccination program with features similar to current recommendations in Canada (i.e., biannual vaccination of those 65 years of age and older and annual vaccination of higher risk individuals less than 65 years of age) showed that such a program can continue to prevent medically attended disease and may be cost-effective in a population with prior immune experience due to infection and/or vaccination.

Results were sensitive to COVID-19 vaccine prices, which are currently unknown for Canada. The price of $43 per dose that was used in the base-case analysis was informed by data on relative price differentials between US CDC public list prices and Canadian negotiated vaccine prices across different vaccine programs. However, given the use of newer vaccine technologies, it remains to be seen if prices for COVID-19 vaccines will follow historical trends. If the Canadian price is similar to that in the US, the ICER for the current program would be $73,400, which exceeds the commonly used cost-effectiveness threshold of $50,000 per QALY. Lack of established vaccine prices remains a challenge for economic modeling to inform vaccination program decisions.

We also found that alignment of vaccine administration with periods of increased SARS-CoV-2 transmission could increase the efficiency of vaccination programs, particularly for those recommended two vaccine doses a year. However, given the current unpredictable nature of the timing of increases in SARS-CoV-2 transmission and considerations of timelines for vaccine strain selection, authorization and supply, and efficiencies of timing COVID-19 vaccination with influenza vaccine administration, the feasibility of changing COVID-19 vaccination program timing may be low. If the epidemiology of COVID-19 becomes more predictable, as is seen for influenza, RSV, and other endemic coronaviruses (6, 7), optimization of vaccination program timing to maximize population protection may become more feasible.

We have focused on evaluating the cost-effectiveness of a COVID-19 vaccination program similar to current recommendations for Canada rather than identifying a vaccination strategy that represents the optimal use of resources. Vaccine coverage estimates for age groups less than 65 years and for the second dose for adults age 65 years and older, which were based on Canadian data (27), were low. The larger number needed to vaccinate to prevent severe outcomes for the younger age group compared to those aged 65 years and older, and the larger ICER in a scenario analysis assuming higher vaccine uptake in the less 65 year age group show that the apparent cost-effectiveness of the overall program may be influenced by lower uptake in groups at lower risk of severe disease. Increased uptake in those at lower risk of severe disease would also increase vaccination program costs. We have conducted a separate analysis looking at cost-effectiveness of vaccination of different population groups to better understand what groups derived the greatest economic benefit from vaccination (30). Similar to other economic evaluations (31, 32), this additional analysis found COVID-19 vaccination programs are most cost-effective when focussed on older age groups at highest risk for severe disease (30). Moving forward, this information may be for ensuring vaccination programs optimize impact in the face of limited budgets.

Our analysis has limitations. The assumption that VE wanes over a period of 7 months was informed by data, but considering the unpredictability of viral evolution, there is a possibility that VE could decline more or less rapidly against circulating strains, which in turn would affect our estimates of cost-effectiveness. The finding that ICERs were higher for the societal perspective than the health system perspective was unexpected, as the inclusion of costs not borne by the health system is generally expected to result in improved cost-effectiveness for economic evaluations of vaccination programs. However, our analysis included productivity losses associated with time and transportation costs to receive the vaccine, resulting in a substantial increase in the costs of the vaccination program when using the societal perspective. Larger ICERs for the societal perspective compared to the health system perspective have been reported previously in an economic evaluation of an immunization program (33) and were similarly attributed to increased immunization program costs. This finding emphasizes the challenges of fully accounting for the societal impacts of disease prevention (34). By focusing on medically attended cases, we are excluding the societal costs of infections that do not require medical care but result in work and school absenteeism and presenteeism, as well as associated caregiver impacts (35–37). Similarly, although we used attributable medical costs for the first year following COVID-19 diagnosis, costs for some may extend beyond one year and we did not capture longer-term decrements in health that may follow COVID-19 disease (38, 39). For these reasons, we expect that we are underestimating the societal costs associated with COVID-19 disease, and this is an important area of future research. Finally, the population groups included in our model focused only on key features of the current Canadian recommendations and do not include all those currently recommended for vaccination. There are several groups 6 months to less than 65 years of age who are currently strongly recommended to receive annual vaccination in NACI recommendations but who do not have chronic medical conditions and were not included in our modelled vaccination strategy. This includes individuals in or from First Nations, Inuit and Métis communities, members of racialized and other equity-denied communities, pregnant women and pregnant people, people who provide essential community services, and residents of long-term care homes and other congregate living settings (9). There is also a discretionary NACI recommendation for average risk individuals 6 months to less than 65 years of age (9), but we did not include this group in our modelled vaccination strategy. In addition, we did not model immunocompromised individuals separately from the higher risk population, although this group is recommended for biannual vaccination (10). The simplification of population groups included in the model was primarily based on data availability; thus our ability to assess cost-effectiveness in these other key population groups was limited.

In summary, our model-based economic evaluation suggests that COVID-19 vaccination programs in Canada remain potentially cost-effective interventions, despite changing epidemiology.

## Data Availability

All data produced in the present work are contained in the manuscript.

## Acknowledgements

The authors are grateful to members of the National Advisory Committee on Immunization COVID-19 Working group for feedback provided during model development and analysis.

## Notes

### Competing Interest Statement

The authors have declared no competing interest.

### Funding Statement

This study did not receive any funding.

